# A Net Benefit Approach for the Optimal Allocation of a COVID-19 Vaccine

**DOI:** 10.1101/2020.11.30.20240986

**Authors:** Erin Kirwin, Ellen Rafferty, Kate Harback, Jeff Round, Christopher McCabe

## Abstract

**OBJECTIVE:** We implement a model-based approach to identify the optimal allocation of a COVID-19 vaccine in the province of Alberta, Canada.

**METHODS:** We develop an epidemiologic model to evaluate allocation strategies defined by age and risk target groups, coverage, effectiveness, and cost of vaccine. The model simulates hypothetical immunization scenarios within a dynamic context, capturing concurrent public health strategies and population behaviour changes.

**RESULTS:** In a scenario with 80% vaccine effectiveness, 40% population coverage, and prioritisation of those over the age of 60 at high-risk of poor outcomes, active cases are reduced by 17% and net monetary benefit increased by $263 million dollars, relative to no vaccine. Concurrent implementation of policies such as school closure and senior contact reductions have similar impacts on incremental net monetary benefit ($352 vs. 292 million, respectively) when there is no prioritisation given to any age or risk group. When older age groups are given priority, the relative benefit of school closures is much larger ($214 vs. 118 million). Results demonstrate that the rank ordering of different prioritisation options varies by prioritisation criteria, vaccine effectiveness and coverage, and concurrently implemented policies.

**CONCLUSIONS:** Our results have three implications: (i) optimal vaccine allocation will depend on the public health policies in place at the time of allocation and the impact of those policies on population behaviour; (ii) outcomes of vaccine allocation policies can be greatly supported with interventions targeting contact reduction in critical sub-populations; and (iii) identification of the optimal strategy depends on which outcomes are prioritised.

## Introduction

Coronavirus disease 2019 (COVID-19) is a severe, novel virus that has spread globally. On 30 January 2020, the World Health Organization (WHO) declared a Public Health Emergency of International Concern and declared a Pandemic on 11 March 2020 [1]. As of 12 March 2021, there have been over 118 million confirmed cases and more than 2.6 million deaths worldwide [1].

While there are several non-pharmaceutical interventions (NPIs) available to reduce transmission among populations, the introduction of effective vaccines is expected to significantly improve decision makers ability to protect health care systems and the economy whilst gaining control of community transmission of COVID-19. Recommended NPIs for COVID-19 mitigation include hand hygiene, mask wearing, physical distancing in various locations, travel restrictions, testing, contact tracing, isolation of cases, and quarantine of close contacts. As vaccines become available, a key challenge for decision makers will be how to concurrently allocate limited vaccine stockpiles, and select optimal combinations of NPIs to mitigate the health risks of COVID-19 while meeting population needs to access the health system for diagnosis and treatment of other conditions, minimising the loss of economic activity, and promoting social well-being.

There are currently more than 200 COVID-19 vaccination products in development, with several in phase III trials, and several have been approved for use in Canada [2, 3]. These vaccines may reduce disease burden by preventing infection in exposed individuals, reducing severity of infection in infected individuals, and/or preventing secondary infections. Within individual health systems, decision makers will need to prioritise access to initially limited vaccine stockpiles [4-6]. Discussions on how to make these prioritisation decisions are well advanced in both the policy and research realms [7-14]. In addition to supply constraints, optimal vaccine allocation will also depend upon population factors such as disease risk, capacity to benefit from vaccine, likelihood of transmission, vaccine effectiveness and vaccine uptake.

The optimal allocation of vaccines will also vary according to the objectives of decision makers. Health economic models of the effectiveness and cost effectiveness of different combinations of NPIs and vaccines can enable more informed vaccine allocation decisions, by capturing the dynamic relationship between vaccine characteristics, allocation strategies, NPIs, and population behaviours. Whilst hundreds of models have been developed internationally to support decision making and public health messaging, only a handful of evaluations of vaccine allocation and prioritization strategies have been published to date [13, 15-18]. Only one of the studies evaluates scenarios with concurrent public health mitigation strategies [13]. None of the studies consider interventions targeting individuals by both age and risk group, with well-defined vaccine allocation policies or health economic outputs, limiting their value to decision makers.

The aim of this paper is to introduce a model-based evaluation of vaccine allocation, analysing strategies defined by age and risk target groups, coverage, and effectiveness, within a dynamic context considering other public health strategies and population behaviour change. Several potential outcomes can be selected as the objective outcome for the optimization problem. Here we consider total number of cases, hospitalizations, and incremental net monetary benefit (NMB). The NMB framework synthesizes multiple model outcomes: mortality, morbidity, and cost of illness. We evaluate hypothetical retrospective scenarios to test a NMB approach to vaccine allocation.

## 1. Methods

We developed the COVID-19 Risk Assessment Model (CRAM), a transmission dynamic ordinary differential equation compartmental susceptible-exposed-infected-recovered (SEIR) model. We calibrated CRAM to the time variant state of public health interventions and human behaviour, using data from Alberta, Canada. To evaluate the risks and outcomes associated with each strategy, scenarios are characterised by a combination of public health interventions that change over time, in comparison to a baseline scenario. Using CRAM we can estimate active cases, hospitalisations, total cost, Quality Adjusted Life Years (QALYs), and NMB. CRAM was programmed in Wolfram Mathematica, version 12.1.

### 2.1 Model Structure

CRAM is stratified into 16 age groups (five-year ranges up to 75+ years). Each of these 16 groups is divided further into two risk groups (high-risk and not high-risk), where risk is based on probability of a poor outcome due to COVID-19. An additional group is used to represent the Long-Term Care (LTC) population, which is made up of a subset of high-risk individuals aged 75 years and older. This results in 33 non-null age-risk groups.

CRAM has five overarching compartments: Susceptible (which includes Susceptible; Susceptible, No Distancing; Vaccine; and Vaccine Failure), Exposed, Infected (composed of Pre-Symptomatic; LTC; Community; Hospital; and Pre-Isolation), Isolation, and Recovered (comprised of Immunised; Recovered; and Dead) (see Figure 1). The model structure was developed to produce outcomes of interest to decision-makers and to leverage the highest quality data available in real-time during the COVID-19 pandemic. Model inputs are provided in Section 2.2.

**Figure 1.**
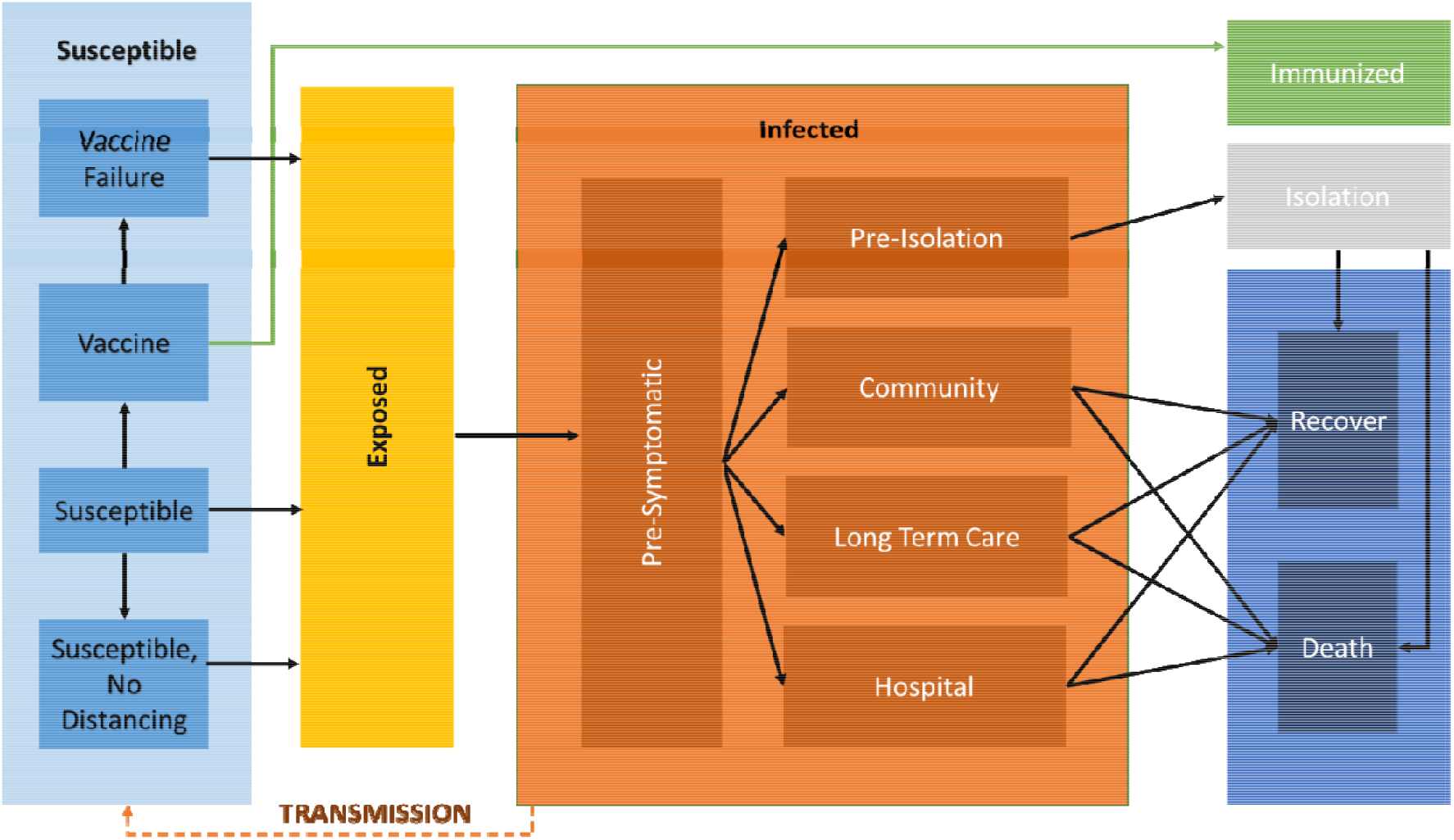
CRAM Structure. **Fig1** ‘Isolation’ compartment is infectious, but does not contribute to the force of infection. Figure intended for colour printing

A fraction of the population is initially allocated to the Exposed compartment, with the remaining population allocated to the Susceptible compartment. Susceptible individuals are vaccinated subject to unique scenarios defined by the following characteristics: (i) program start date, (ii) number of doses available, (iii) the daily immunization delivery capacity and (iv) age and risk group prioritisation policy. Vaccine prioritisation policies allow access to different age and risk groups on different dates. Once vaccinated, after the period required to develop antibodies, individuals move either to the Immunised compartment (with no probability of infection or transmission), or to the Vaccine Failure compartment (with equal probability of infection and transmission to others in the susceptible compartment) as a function of the vaccine effectiveness. Vaccine Failures remain susceptible.

Susceptible individuals are exposed to the virus as a function of the force of infection which is determined by the transmissibility of the virus, seasonal variations in transmission, number of persons infected, and rate of contact between individuals across age groups. Susceptible individuals exposed to the virus move to the Exposed compartment and following the latent period move from the Exposed to Infected.

Depending on public health guidance, some individuals may move from the Susceptible compartment to the Susceptible, No Distancing compartment which allows for higher rates of contact. For example, children who are not high-risk move to the Susceptible, No Distancing compartment to reflect the higher rates of contact from in-person education.

On entry to the Infected compartment, individuals are Pre-Symptomatic. That is, cases in the community that are shedding virus but not experiencing or displaying symptoms. At the end of the pre-symptomatic period, individuals are allocated to Community or Hospital compartments, dependent on their age and risk group. If a case isolation policy is in place, a subset of the Community compartment will move to the Pre-Isolation and subsequently the Isolation compartments, where they remain for the remaining infectious period. All cases contribute to the force of infection prior to moving to the Isolation compartment, but individuals in the Isolation compartment do not contribute to the force of infection. The Isolation compartment represents the isolation of cases but not the isolation of close contacts.

Entry into LTC and Hospital varies by age group, with individuals from the LTC age-risk group moving from the pre-symptomatic to the infected LTC compartment. Individuals move from the LTC compartment at the end of the infectious period to either the Recovered or Dead compartments. Entry into the Hospital compartment is a function of age and risk, fit by an exponential regression on admissions by age, and adjusted for risk by an odds ratio. The Hospital compartment includes COVID inpatient cases and those admitted to intensive care units (ICUs). At the end of either the infectious period or the hospital length of stay (LOS), individuals move from the Community, LTC or Hospital compartment to either the Recovered, or Dead compartment, at a proportion dependent upon their risk group.

We used a synthetic contact matrix disaggregated into four locations: home, workplace, school, and other, where other comprises all remaining possible locations [19]. Disaggregating the total contact matrix by location allows for adjustments in contact rates reflecting policy interventions. For example, we can apply work-based interventions to specific age groups by adjusting the work contact matrix at various points in time. The total contact matrix is generated by summing the contacts for each age group across each of the matrices. Contact reductions for seniors aged 75 years and older and for high-risk individuals are applied directly to the total contact matrix. Outcomes (cases, hospitalizations, deaths) are tabulated from 01 October, 2020 to 06 February, 2021, spanning 92 days. The Electronic Supplementary Material (ESM) reports model equations and assumptions.

### 2.2 Model Inputs

CRAM was parameterised wherever possible with data from Alberta, including estimates for the total population by age group and risk of severe outcome, hospitalisation rate by age group, and LOS for hospitalised cases. Alberta’s population is the youngest of all Canadian provinces, making age distribution important to capture in the model [20]. Odds ratios for the risk of hospitalisation were estimated by risk group using logistic regression. Active cases in the model include both diagnosed and undiagnosed cases, fitted using seroprevalence data provided by Alberta Health. Details of all model inputs are provided in Table 1.

**Table 1.**
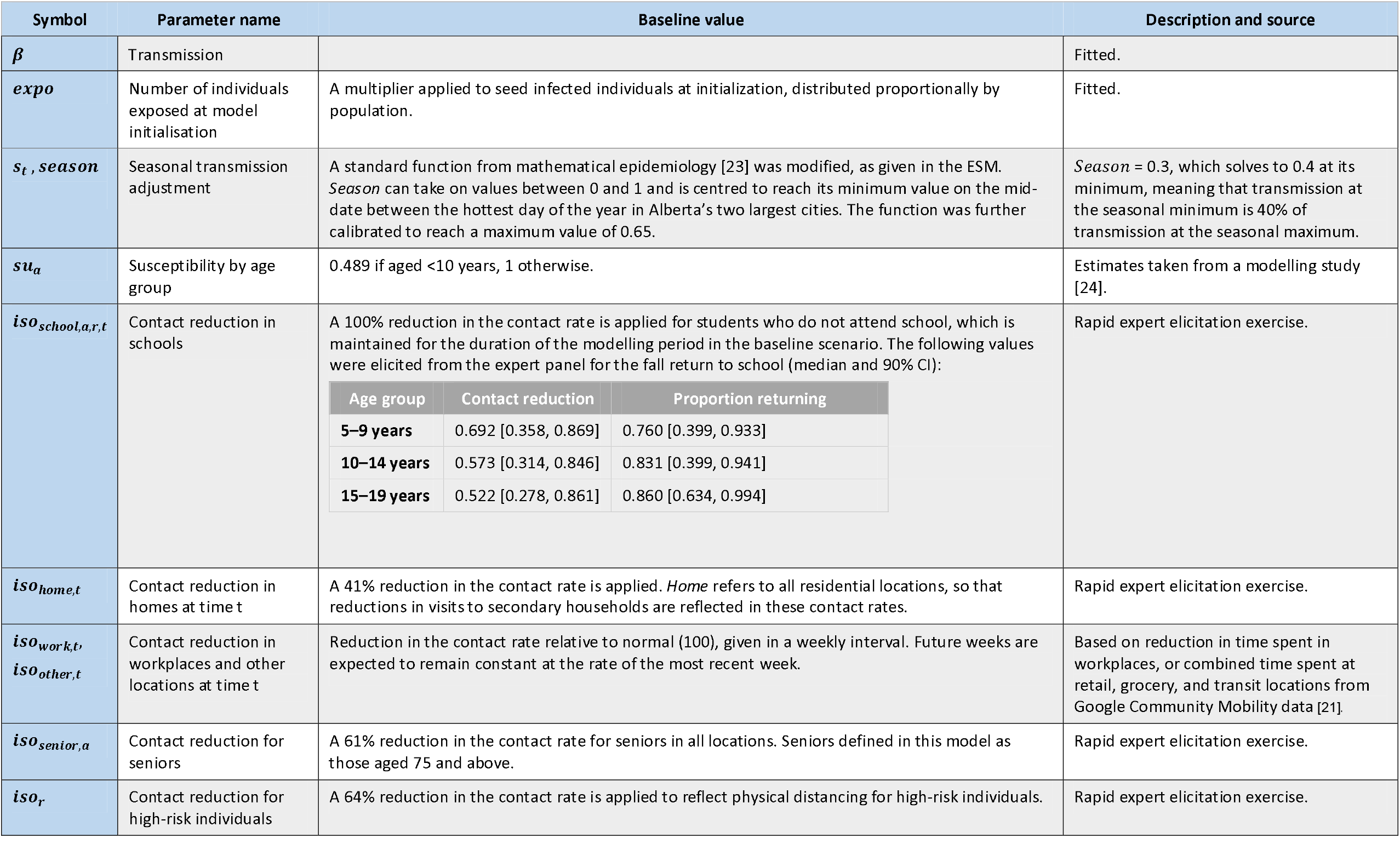
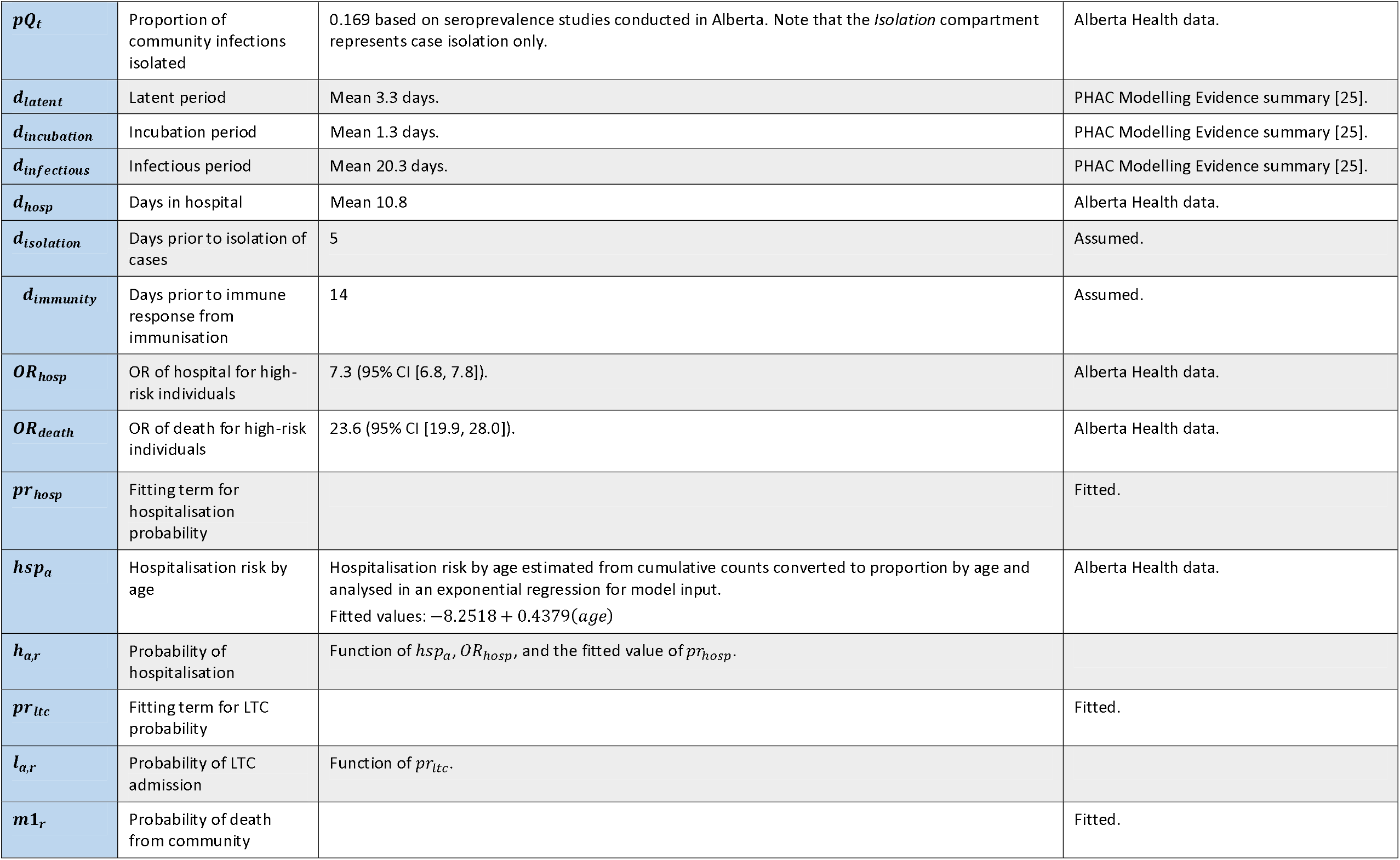
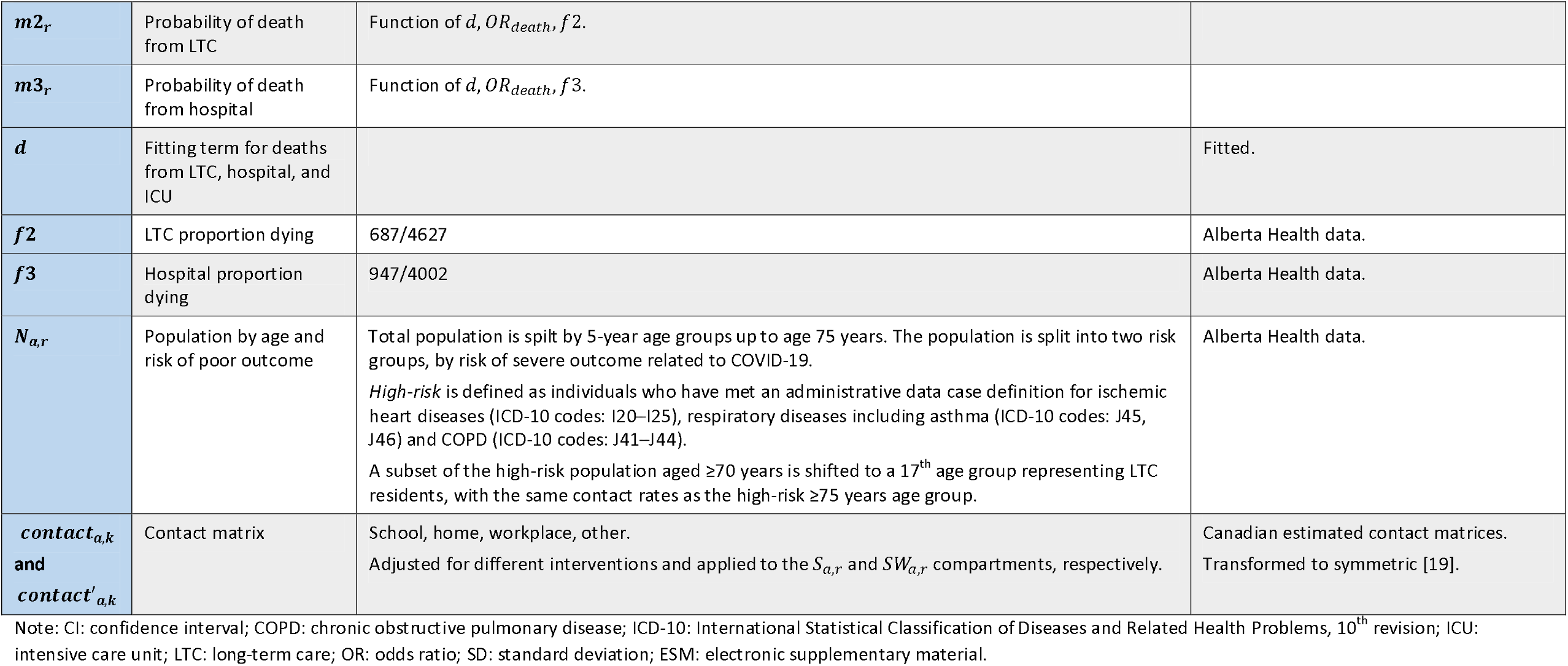
Epidemiologic Parameters.

Data from the Google Community Mobility report [21] were used as key inputs for contact reductions in workplaces and other locations. A rapid expert elicitation exercise was conducted to obtain estimates for unknown model parameters. We adapted elicitation methods and training materials from the Sheffield Elicitation Framework (SHELF) [22] to undertake rapid elicitations for the reduction in contacts for seniors aged 75 years and older, high-risk individuals, reduction in contacts in home locations, and expected return-to-school rates and in-school contact reductions by age group. Detailed results of the elicitations are reported in the ESM.

### 2.37 Economic Inputs

The study is conducted from a Canadian health system payer perspective. All costs are converted to 2020 Canadian dollars, and discounted at an annual rate of 1.5%, according to CADTH guidelines [26]. Cost inputs include hospitalisation, incremental LTC costs for infection prevention measures, and the cost of purchasing and administering a vaccine. Health utility losses include disutility from infection, hospitalisations and LTC infections (during stay and following discharge), and deaths. We assessed outcomes using the NMB framework [27]. We apply the health opportunity costs estimated for Canada [28], using the value of $30,000 per QALY.

Evidence related to health utility for COVID-19 is currently limited but rapidly emerging. [29] Given the lack of evidence at the time of writing, we identified utility impacts of similar conditions. We estimated disutility for community COVID-19 infections from an EQ-5D survey of individuals with lab confirmed influenza B infections, stratified by age group [30]. Because hospitalised COVID-19 cases are often diagnosed with pneumonia, we used disutility estimates for viral pneumonia inpatients [31] to estimate in-facility treatment utility losses, as well as utility losses for the year following facility discharge. We estimated QALY losses from premature death using the life table approach developed by Briggs et al. [32], adjusting for baseline utility and discounting. Costs related to community infections are not included. We estimated incremental direct costs associated with COVID-19 hospitalisation based on published Alberta per-diem costs, as well as the incremental cost of infection prevention and control for LTC cases. We also estimate utility decrements for ‘chronic’ or ‘long’ infections (ESM). Cost and utility decrement estimates are given in Table 2. Estimates are applied to the relevant compartments, and the resulting incremental NMB (iNMB) is estimated for each policy scenario.

**Table 2.**
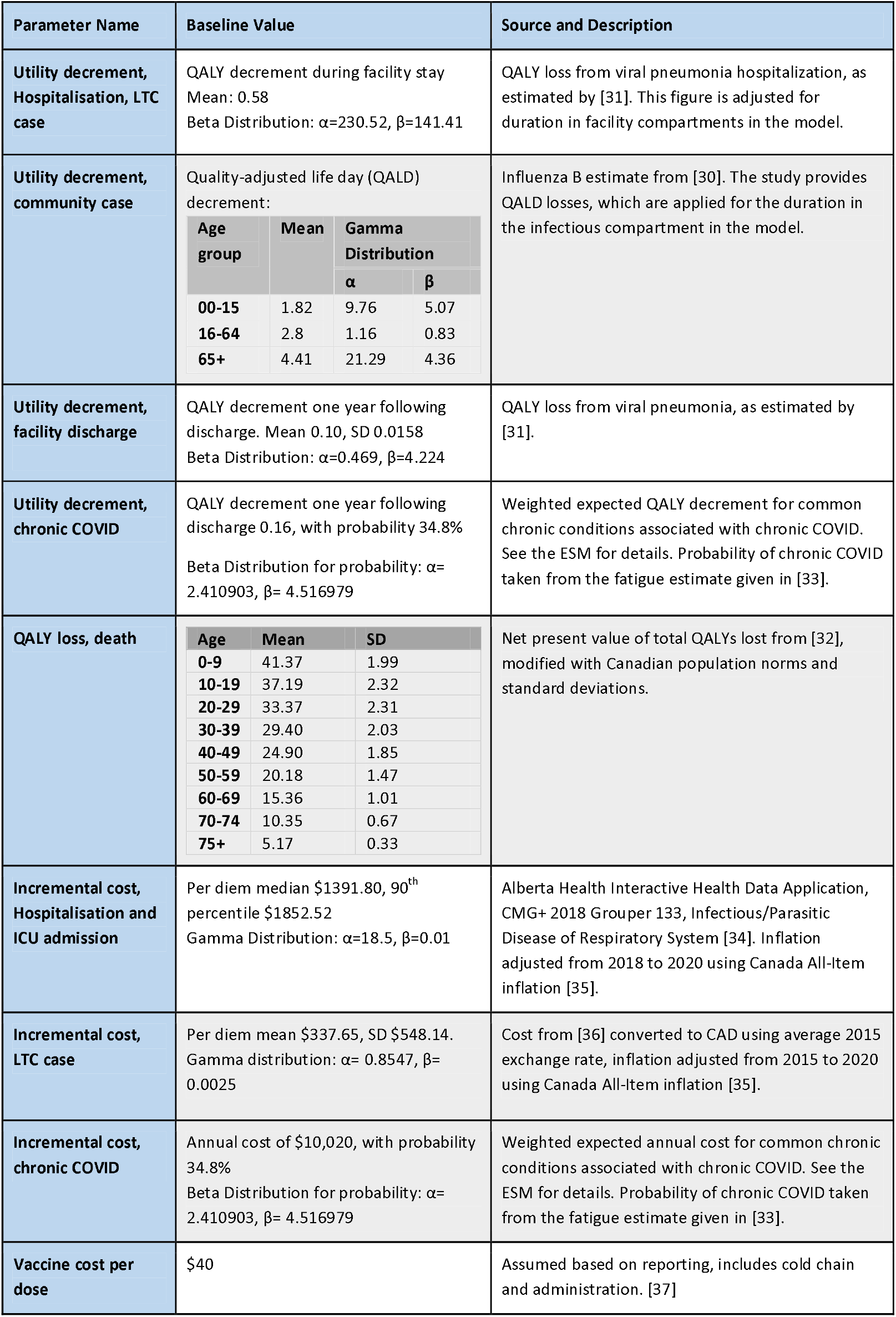
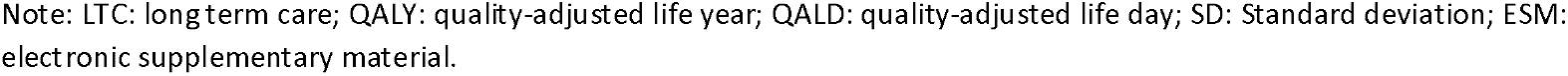
Cost and Utility Parameters.

### 2.4 Model Fitting

We fit five parameters: virus transmissibility, the probability of hospitalisation, infections in LTC, and the probability of death from either facility or community infections. Using a deterministic version of the model defined with mean values of each input parameter we fit observed surveillance data for daily hospital census counts, as well as cumulative daily death and LTC counts from 01 October 2020, to 06 February 2021. Given the interdependence in the fitting parameters, we used simultaneous fitting methods by optimizing model fit over a Latin hypercube generated from plausible values for each parameter. Optimal parameter values were identified when the sum of the normalised difference in model and observed outcomes was minimised.

### 2.5 Outputs

CRAM samples from the probability distributions of model inputs, as either the best fitting 10% of simulations from the model fitting, or as estimated from the parameter source (Tables 1 & 2), to account for outcome uncertainty over 200 iterations, and then generates model outputs from the sampled values. This number of iterations was selected after comparing output stability across different numbers of iterations, for details, see the ESM. Outputs include cumulative estimates for the mean number of active cases, total infections, and hospitalisations, with credibility intervals to present uncertainty over simulations. The outputs include expected NMB, so that comparisons without the use of ratios can be made [27]. For each of the vaccine scenarios, iNMB is estimated relative to the baseline with no vaccine.

### 2.6 Scenario Analysis

In the reference scenario (1), no vaccine is available, and all contact matrixes and time variant inputs are held constant at the last observed value from model fitting. Specifically, we maintained contact reductions at the values determined through the expert elicitation for ‘home’ locations, as well as for seniors and high-risk individuals, and a fall return to school. Moreover, we kept the final observation of reduction in time spent in workplaces and ‘other’ locations from the Google Community Mobility reports consistent over the model run. In all scenarios, the vaccine is ‘sterilizing’, meaning that effective vaccine protects individuals from infection.

A range of NPIs can be estimated by CRAM. Policies which impact contact rates in the home, workplace, school and other locations for different age groups can be simulated through reductions on the corresponding contact matrix. Similarly, broad age-based interventions can be applied to the total contact matrix. In this paper, we considered school closure via a complete reduction in school-based contacts, and policies with increased physical distancing for seniors by reducing contact rates for those aged 75 and older by 50%.

Vaccine scenarios represent different vaccine allocation strategies, as well as various vaccine characteristics (i.e., effectiveness and coverage), as described in Table 3. Vaccine scenarios are characterised by the maximum number of doses that can be delivered per day, the start date of the vaccine program, the definition of the priority group, the interval (if any) between access for the priority group and the general population, the effectiveness of the vaccine, and vaccine coverage as a proportion of the total population. The estimates of vaccine effectiveness used in the analysis do not directly align to any of the currently approved vaccine products in Canada or elsewhere, nor does the price or timing of allocation align to the initial approval or distribution of licensed vaccines.

**Table 3.** Vaccine Scenarios.

| Scenario<br>(Name, Reference<br>Number) |  | Prioritization Policy | Vaccine Effectiveness | Population Dose<br>Coverage |
| --- | --- | --- | --- | --- |
| No Vaccine | 1 | No Vaccine | N/A | N/A |
| Vaccine<br>Baseline | 2 | Age + Risk | 80% | 40% |
|  | 2a, 2b | Same as 2, but with school closures (2a) or decreased contact rates for seniors (2b), with varying prioritization policies. |  |  |
| Vaccine<br>Coverage<br>Comparison | 3 | Age + Risk | 80% | 20% |
|  | 4 |  |  | 70% |
| Vaccine<br>Effectiveness<br>Comparison | 5 | Age + Risk | 40% | 40% |
|  | 6 |  | 60% |  |
|  | 7 |  | 60% above 60 years old, 80% otherwise |  |
| Vaccine<br>Priority Group<br>Comparison | 8 | No Priority | 80% | 40% |
|  | 9 | Age Only |  |  |
|  | 10 | Risk Only |  |  |
| Low<br>Effectiveness<br>and Coverage | 11 | Age + Risk | 40% | 20% |
|  | 11a, 11b | Same as 11, but with school closures (11a) or decreased contact rates for seniors (11b), with varying prioritization policies. |  |  |

Note: Age + Risk = high risk population, age > 55, Age Only = age >55, Risk Only = high risk population.

If the number of individuals eligible for immunisation exceeded the maximum number of doses that could be delivered on a given day, the maximum number of doses were pro-rated across each eligible age and risk group daily. This means that a group representing 5% of the eligible population would receive 5% of the total number of doses available on a given day.

To examine the impact of variations in vaccine effectiveness, we included scenario 7, where effectiveness varies by age. It is assumed that the vaccine is 60% effective over the age of 60, and 80% effective otherwise. In all other scenarios, effectiveness is constant across age groups. The scenario with 80% effectiveness approximates preliminary results from three phase three trials which have reported efficacy ranging from approximately 70 to 94% [38-40].

In each of the scenarios (1-11), we assumed a simple 30-day interval in access for the prioritized population groups and the general population, starting on 01 November 2020. Depending on the prioritization policy, this could lead to a gap of days during which no immunization would be provided. Extrapolating from the maximum estimate of vaccines per day in Alberta from [41], we assumed that up to 80,000 individuals can be immunised daily. Each of the scenario comparisons implicitly contains the baseline scenario (2) with age and risk-based prioritisation, 80% effectiveness, and 40% coverage, as a common reference point across scenarios. Scenarios that prioritise based on age give first access to those aged 55 and over, while programs that prioritise high-risk individuals prioritise all high-risk individuals irrespective of age.

Scenario 11 is defined with low vaccine effectiveness (40%) and coverage (20%). For both scenario 11 and the baseline scenario (2), we modelled additional public health measures, including a 50% reduction in contacts for seniors, and complete school closures (each policy implemented on 01 October 2020). For both sub-scenarios (school closures and seniors contact reduction), we assumed no additional costs or QALY losses. These simulations compare the impact vaccine policies in combination with other mitigation strategies, albeit for a small range of potential policy combinations.

## 3. Results

We used the CRAM to produce outputs for each vaccine scenario. For all scenarios, mean values, and either standard deviation or a 90% credibility interval are presented. In the no vaccine scenario, total cases were shown to remain relatively low in the initial period, but increase rapidly over time. In the vaccine baseline scenario (scenario 2), active cases are reduced by 17%. The impact of population vaccine coverage on outcomes demonstrates improved outcomes with greater population vaccine coverage, as expected. When coverage is 70%, cases are significantly reduced by 38%. Interestingly, with 70% coverage, iNMB decreases relative to baseline (Table 4, $216 vs 263 million). This indicates decreasing returns to scale, related to approaching herd immunity. The impact of vaccine effectiveness on outcomes behaved in the expected manner, where NMB increases alongside effectiveness. With 40% effectiveness, the availability of a vaccine reduces cumulative cases by 10%, while with 60% effectiveness, cumulative cases decrease by 14%. These results are summarised in Table 4 and Figure 2.

**Figure 2.**
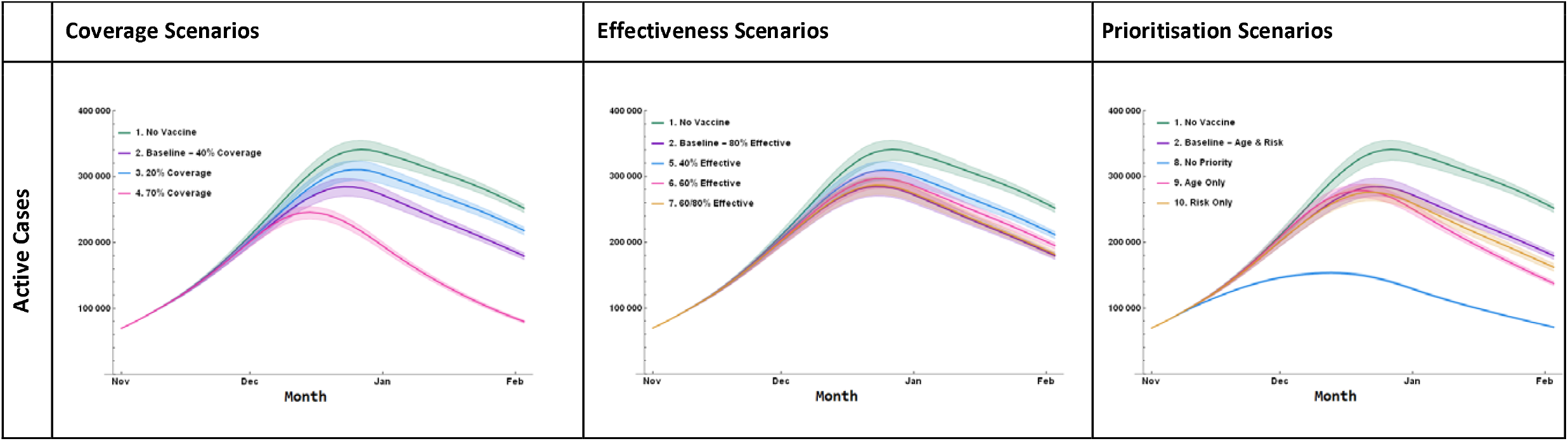
Scenario Results - Active Cases. **Fig2** For scenario definitions, see main text

Vaccine prioritisation strategies demonstrated dynamic results. Prioritizing both high-risk and individuals over the age of 55 had the smallest impact on infections and produced the second lowest value for iNMB. Equal prioritisation to all age and risk groups minimised cumulative cases and had the greatest iNMB value. Prioritisation based on risk levels alone had the second highest iNMB value. Prioritization by age group had the second greatest reduction in cases, but the lowest reduction in iNMB relative to the scenario with no vaccine. The variability in results demonstrates the need for decision makers to clearly identify their objective when determining vaccine allocation strategies.

The results of the concurrent NPI scenarios (Table 5) demonstrate several important relationships. First, the relative ranking of each prioritisation scenario varied depending on the outcome of interest. For example, in the worst case scenarios 11a and 11b, risk based prioritization had the second best iNMB and number of hospitalizations, but the second worst number of cases (Figure 3, Table 5).

**Figure 3.**
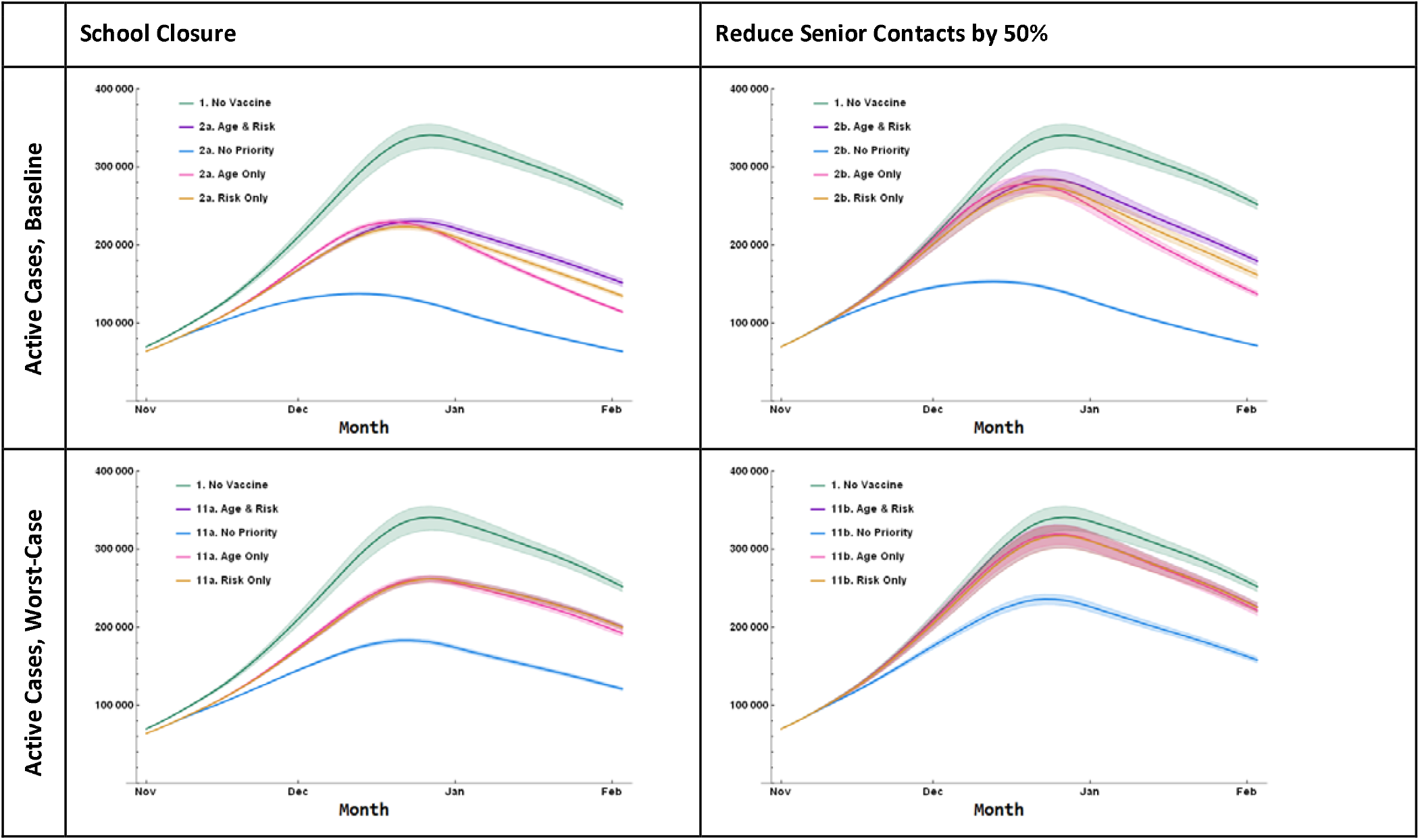
Concurrent Non-Pharmaceutical Intervention Results – Active Cases. **Fig3** For scenario definitions, see main text

Second, the impact of prioritisation strategies varies greatly depending on concurrent public health interventions. School closures and senior contact reductions have similar impacts on iNMB when there is no prioritisation given to any age or risk group (Table 5, iNMB 352 vs. 292 million, respectively, ratio 1.2:1), but when older groups are given priority, the benefit of school closures is much larger than reducing contacts for seniors (Table 5, iNMB 214 vs. 118 million, respectively, ratio 1.8:1).

Third, we found that the relative ranking of the prioritisation scenarios changed dynamically between the baseline scenario (Table 4) and the concurrent NPI scenarios (Table 5). In all scenarios, giving equal priority to all age and risk groups maximised iNMB, and minimized cases and hospitalizations in all concurrent NPI scenarios. Conversely, prioritizing older and high-risk individuals had the greatest reduction in hospitalizations in the baseline scenario with no concurrent NPIs.

### 3.1 Coverage, Effectiveness, and Prioritisation Scenario Results

**Table 4:** Scenario Results.

| Scenario<br>(Reference Number, Name) |  | Cumulative Cases<br>Mean (SD) | Cumulative<br>Hospitalisations<br>Mean (SD) | NMB, Millions<br>Mean (SD) | iNMB*, Millions<br>Mean (SD) |
| --- | --- | --- | --- | --- | --- |
| **Reference Scenarios** |  |  |  |  |  |
| **1** | **No Vaccine** | 12.4 (0.2) | 6380 (238) | -980 (112) | n/a |
| **2** | **Vaccine Baseline<sup>†</sup>** | **10.3 (0.2)** | **3419 (112)** | **-718 (91)** | **263 (130)** |
| **Coverage Scenarios** |  |  |  |  |  |
| **3** | **Coverage 20%** | 11.3 (0.2) | 4580 (162) | -805 (100) | 175 (132) |
| **4** | **Coverage 70%** | **7.7 (0.1)** | **2975 (92)** | **-764 (77)** | **216 (130)** |
| **Effectiveness Scenarios** |  |  |  |  |  |
| **5** | **Effectiveness 40%** | 11.2 (0.2) | 4760 (168) | -902 (100) | 78 (140) |
| **6** | **Effectiveness 60%** | 10.7 (0.2) | 4061 (139) | -807 (95) | 174 (142) |
| **7** | **Effectiveness 60% age 60 and above, 80% otherwise** | **10.4 (0.2)** | **3729 (125)** | **-752 (91)** | **228 (138)** |
| **Prioritisation Scenarios** |  |  |  |  |  |
| **2** | **Age and Risk (Vaccine Baseline)<sup>†</sup>** | 10.3 (0.2) | **3419 (112)** | -718 (91) | 263 (130) |
| **8** | **None** | **5.9 (0.0)** | 3432 (103) | **-692 (60)** | **288 (124)** |
| **9** | **Age Only** | 9.4 (0.2) | 4277 (147) | -865 (93) | 115 (142) |
| **10** | **Risk Only** | 9.8 (0.2) | 3470 (116) | -706 (88) | 274 (136) |

Note: SD = Standard Deviation, NMB = Net Monetary Benefit, iNMB = Incremental Net Benefit, N/A = Not Applicable. Prioritisation Scenarios are defined as follows: None = equal priority given to all age and risk groups, Age = priority given to those aged 55 and over, Risk = priority given to high-risk individuals, Age and Risk = prioritisation for who are high-risk or older than 55. Maximum doses per day is assumed to be 80,000, with access starting on October 15, 2020 for the priority group, and 30 days later for everyone else. \*For all scenarios, iNMB is estimated relative to the scenario with no vaccine. Within each scenario group, the optimal policy is bolded. The dagger (†) denotes the same scenario repeated in the table for ease of reference.

### 3.2 Low Effectiveness and Coverage Scenarios: Concurrent Public Health Responses

**Table 5.** Concurrent Non-Pharmaceutical Intervention Results.

| Scenario and Concurrent public health interventions | Prioritisation | Cumulative Cases<br>Mean (SD), Rank (1-4) |  | Cumulative Hospitalisations<br>Mean (SD), Rank (1-4) |  | NMB, Millions<br>Mean (SD), Rank (1-4) |  | iNMB*, Millions<br>Mean (SD), Rank (1-4) |  |
| --- | --- | --- | --- | --- | --- | --- | --- | --- | --- |
| 2a: Baseline, School Closure | Age and Risk | 8.6 (0.1) | 4 | 3061 (92) | 2 | -646 (76) | 3 | 334 (135) | 3 |
|  | None | 5.2 (0.0) | 1 | 3006 (87) | 1 | -628 (53) | 1 | 352 (120) | 1 |
|  | Age Only | 7.9 (0.1) | 2 | 3738 (116) | 4 | -767 (77) | 4 | 214 (138) | 4 |
|  | Risk Only | 8.2 (0.1) | 3 | 3086 (91) | 3 | -633 (73) | 2 | 348 (134) | 2 |
| 2b: Baseline, Reduce senior contacts 50% | Age and Risk | 10.3 (0.2) | 4 | 3366 (110) | 2 | -715 (90) | 3 | 266 (132) | 3 |
|  | None | 5.9 (0.0) | 1 | 3364 (101) | 1 | -688 (60) | 1 | 292 (125) | 1 |
|  | Age Only | 9.4 (0.2) | 2 | 4227 (144) | 4 | -862 (92) | 4 | 118 (149) | 4 |
|  | Risk Only | 9.8 (0.2) | 3 | 3412 (112) | 3 | -703 (88) | 2 | 278 (129) | 2 |
| 11a: Worst Case, School Closure | Age and Risk | 9.9 (0.1) | 4 | 4480 (144) | 3 | -765 (87) | 3 | 215 (131) | 3 |
|  | None | 7.2 (0.1) | 1 | 4031 (126) | 1 | -701 (67) | 1 | 280 (119) | 1 |
|  | Age Only | 9.8 (0.1) | 2 | 4636 (150) | 4 | -809 (88) | 4 | 171 (128) | 4 |
|  | Risk Only | 9.9 (0.1) | 3 | 4432 (142) | 2 | -748 (86) | 2 | 232 (139) | 2 |
| 11b: Baseline, Reduce senior contacts 50% | Age and Risk | 11.6 (0.2) | 4 | 4909 (177) | 3 | -858 (102) | 3 | 122 (131) | 3 |
|  | None | 9.0 (0.1) | 1 | 4735 (160) | 1 | -820 (84) | 1 | 161 (131) | 1 |
|  | Age Only | 11.6 (0.2) | 2 | 5157 (183) | 4 | -915 (106) | 4 | 66 (149) | 4 |
|  | Risk Only | 11.6 (0.2) | 3 | 4856 (174) | 2 | -839 (102) | 2 | 142 (136) | 2 |

Note: SD = Standard Deviation, NMB = Net Monetary Benefit, iNMB = Incremental Net Benefit. Prioritisation Scenarios are defined as follows: None = equal priority given to all age and risk groups, Age = priority given to those aged 55 and over, Risk = priority given to high-risk individuals, Age and Risk = prioritisation for who are high-risk or older than 55. Maximum doses per day is assumed to be 80,000, with access starting on October 15, 2020 for the priority group, and 30 days later for everyone else. \*For all scenarios, iNMB is estimated relative to the scenario with no vaccine. Within each scenario group, rank 1 gives the optimum policy, whereas rank 4 gives the worst, ranking is based on unrounded results.

## 4. Discussion

Our results demonstrate the conflicts faced by decision makers when determining which policy objectives to pursue. In addition to the expected result that the choice of optimal vaccine allocation strategy is dependent on the primary outcome of interest, we also show the importance of considering time-variant epidemiological conditions, NPIs in place while a vaccine stockpile is allocated, vaccine coverage and effectiveness, population prioritisation policies, and timing of vaccine availability.

Cumulative cases and iNMB vary greatly depending upon which concurrent strategies are in place. The optimum vaccine allocation strategy is sensitive to whether policies target morbidity reductions (cases, hospitalizations) or NMB outcomes. The choice over which outcomes to optimize is a reflection of values, and/or of specific concerns (such as hospital capacity) at the time of decision making. Depending on contact patterns and case burden, increases in cases do not imply equivalent marginal increases in hospitalizations, and vice-versa. Similarly, outcomes such as hospitalizations have different impacts by age and risk group, which are not captured in count-based measures. The NMB measure combines the QALY losses and health system costs related to multiple outcomes (cases, hospitalizations, deaths).

With the exception of Chen et al. [13], other studies do not consider concurrent or future public health policies/ and or behavioural factors. We consider this an important oversight, as vaccine roll outs will take place in the context of other public health interventions and variation in behavioural responses. Ignoring this dynamic would risk misallocating the limited and highly valuable vaccine supply. Similarly, very few other studies have examined allocation by age and risk, [13,15,17] which are key factors in vaccine allocation discussions.

Our study applies a health system perspective and incurs immunisation costs, whereas a societal perspective is expected to broadly increase NMB in immunisation scenarios relative to a health system perspective as broader social, economic, and health impacts are considered. For example, the societal cost of school closures are not captured in model results. The model is run for a short time frame such that the full benefits of immunisation are not fully realised, and important dynamics such as waning immunity are unknown and not considered in this analysis. Despite these limitations, the large NMB loss estimates for all scenarios demonstrate the magnitude of the harm caused by the COVID-19 pandemic, and the relative impact of vaccination on NMB in the short term.

Some results from our analysis are unexpected and warrant further explanation. The impact of a lower vaccine effectiveness in those aged 60 and over appears relatively small. This is likely due to the expectation that physical distancing is maintained at a higher level for seniors throughout the period of analysis. If public health guidance or population attitudes shift such that seniors have increasing contact rates, lower effectiveness for older age groups would have a greater impact.

The results for the ‘no prioritization’ strategy is non-intuitive. While age and risk groups that are older or high-risk have a greater risk of hospitalisation or death from COVID, these outcomes interact dynamically with greater transmission, driven by higher rates of contact, in young and low risk age groups. With increasing evidence of COVID-19 vaccines reducing the probability of viral transmission [42, 40], there is an argument that immunizing those with greater rates of contact will confer greater population level benefits [43]. This dynamic may also be due to relatively high coverage in the scenarios evaluated. These effects may not be as strong with coverage lower than 40%, as evaluated in our baseline scenarios. With sufficient evidence to support sterilizing immunity, this is an important finding as the implication will be that individuals who cannot avoid contact (due to their employment, socioeconomic status, or living situation) should be prioritized for immunization to minimize onwards transmission and increase protection at a population level.

We have modelled ‘all or nothing’ vaccine effectiveness. Other authors have shown that evaluating ‘leaky’ effectiveness impacted upon the dynamics of the population impact [17]. This would not change the importance of the dynamic relationships our analysis elucidates but would impact upon the relative magnitude of different policy strategies. Future analyses with CRAM could incorporate evidence-based vaccine effectiveness and the identification of critical coverage thresholds needed to achieve herd immunity.

Our analysis assumes only susceptible individuals are immunized and does not consider the potential role of serological testing for guiding vaccination strategies [17]. The assumption that only susceptible individuals are tested implies that an accurate method of case identification is in place, and more realistic evaluations intended to inform policies must consider the likely population distribution of recovered cases, to ensure realistic modelling of immunizing both susceptible and recovered individuals.

Furthermore, evidence of marginal vaccine effectiveness on recovered individuals would be required. As stated in the introduction, the aim of this paper is to develop a model-based approach to vaccine allocation, analysing strategies defined by age and risk target groups, coverage, and effectiveness, within a dynamic context considering NPIs and population behaviour change. To this end, we evaluated intentionally hypothetical scenarios to avoid direct application of our results rather than our approach to vaccine allocation. Analyses designed to inform decision making must reflect the context within decisions are being made: current epidemiology, the timing of vaccine allocation, vaccine supply, delivery capacity, product effectiveness, proposed dose intervals, inter-dose effectiveness, concurrent NPIs, and population behaviour.

Our results have three key policy implications: (i) that optimal vaccine allocation will depend on the public health policies, epidemiology, and human behaviours in place at the time of allocation; (ii) the effectiveness and efficiency of vaccine allocation policies can be greatly enhanced by co-interventions that target contact reduction in critical sub-populations; and (iii) the identification of the optimal strategy depends on which outcomes are prioritised.

Future analyses conducted to inform policy directly should reflect these findings. It is critical to identify which outcomes are prioritized in vaccine allocation. Decision makers must not only consider the interventions and behaviours in place at the time of allocation, but also those which could be in place in the future. Whereas our paper evaluates changes to NPIs in separate scenarios, optimal allocation will depend on public health policy, epidemiology, and human behaviour. This creates a dynamic decision-making environment where different combinations and degrees of implementation should be compared for all NPIs. Similarly, it is likely that real-world decision problems will involve the allocation of multiple vaccines, targeting different segments of the population with different availability over time. The full suite of policy options (both NPIs and vaccine allocation options) and combinations thereof must be examined to identify an optimal solution.

Public health policies that can be implemented alongside vaccine allocation must be considered within a clear framework reflecting social values and equity. For example, in our model NMB is improved relative to baseline when seniors continue to reduce their number of contacts, and if vaccines are allocated without prioritizing older and high-risk population groups. However, this would mean that the people with the greatest risk of disease and poor outcomes would also bear the highest burden through additional social isolation. Therefore, the suite of public health policy policies to be implemented alongside the vaccine should be determined prior to dynamic policy optimisation [8].

## Data Availability

All data sources for the model are listed directly in the manuscript.

## Acknowledgments

The authors acknowledge Dr. Marie Varughese and Li Huang for providing epidemiologic data for model input. We also thank Joana Gomes da Costa for assistance in identifying information related to chronic COVID. We acknowledge earlier work on influenza modelling in Alberta, the Pandemic Risk Assessment Model, with Dr. Douglas Dover.

## Declarations

### Availability of data and material

All data sources documented.

### Code availability

Available from authors upon request.

### Author Contributions

All authors were involved in the conception of the study. EK designed, programmed, and calibrated the model. EK, ER and KH contributed to the collection of data. EK wrote the first draft of the manuscript with ER, KH, JR and CM critically revising for important intellectual content. All authors read and approved the final manuscript.

