## Supplementary Material for "A Net Benefit Approach for the Optimal Allocation of a COVID-19 Vaccine"

**Journal: PharmacoEconomics**

**Electronic Supplementary Material**

#1200, 10405 Jasper Avenue

Edmonton, AB, Canada T5J 3N4

### 1 Model Details

#### 1.1 Equations

The full set of equations used to describe CRAM is provided in this appendix. Each compartment is divided into 17 age groups and two risk groups. The 34 (33 non-null) sub-compartments are indexed by *a,r*, where *a* is the age group and *r* is the risk group. The population size of each group is given by $N_{a,r}$ and a compartment sub-indexed by $N_{a,\cdot}$ represents the sum of both risk groups for one age group.

$$S'_{a,r} = -\beta * su_a * s_t * iso_r * \sum_{k=1}^{17} \frac{contact_{a,k}}{N_{k,\cdot}} [IPS_{k,\cdot} + IC_{k,\cdot} + IQH_{k,\cdot} + IL_{k,\cdot} + IH_{k,\cdot}] * S_{a,r} - w_{a,r,t} * S_{a,r} - v_{a,r,t} * S_{a,r}$$

$$SW'_{a,r} = w_{a,r,t} * S_{a,r} - \beta * su_a * s_t * \sum_{k=1}^{17} \frac{contact'_{a,k}}{N_{k,\cdot}} [IPS_{k,\cdot} + IC_{k,\cdot} + IQH_{k,\cdot} + IL_{k,\cdot} + IH_{k,\cdot}] * SW_{a,r} - v_{a,r,t} * SW_{a,r}$$

$$V'_{a,r} = v_{a,r,t} * S_{a,r} + v_{a,r,t} * SW_{a,r} - \beta * su_a * s_t * iso_r * \sum_{k=1}^{17} \frac{contact_{a,k}}{N_{k,\cdot}} [IPS_{k,\cdot} + IC_{k,\cdot} + IQH_{k,\cdot} + IL_{k,\cdot} + IH_{k,\cdot}] * V_{a,r} - d_{immune} * V_{a,r}$$

$$VF'_{a,r} = (1 - VE) * d_{immune} * V_{a,r} - \beta * su_a * s_t * iso_r * \sum_{k=1}^{17} \frac{contact_{a,k}}{N_{k,\cdot}} [IPS_{k,\cdot} + IC_{k,\cdot} + IQH_{k,\cdot} + IL_{k,\cdot} + IH_{k,\cdot}]$$

$$E'_{a,r} = \beta * su_a * s_t * iso_r * \sum_{k=1}^{17} \frac{contact_{a,k}}{N_{k,\cdot}} [IPS_{k,\cdot} + IC_{k,\cdot} + IQH_{k,\cdot} + IL_{k,\cdot} + IH_{k,\cdot}] * (S_{a,r} + V_{a,r} + VF_{a,r}) + \beta * su_a * s_t * \sum_{k=1}^{17} \frac{contact'_{a,k}}{N_{k,\cdot}} [IPS_{k,\cdot} + IC_{k,\cdot} + IQH_{k,\cdot} + IL_{k,\cdot} + IH_{k,\cdot}] * SW_{a,r} - d_{latent} * E_{a,r}$$

$$IPS'_{a,r} = d_{latent} * E_{a,r} - d_{incubation} * IPS_{a,r}$$

$$IC'_{a,r} = (1 - h_{a,r} - l_{a,r}) * (1 - pQ_t) * d_{incubation} * IPS_{a,r} - d_{infectious} * IC_{a,r}$$

$$IQH'_{a,r} = (1 - h_{a,r} - l_{a,r}) * (pQ_t) * d_{incubation} * IPS_{a,r} - d_{isolation} * IQH_{a,r}$$

$$IQ'_{a,r} = d_{isolation} * IQH_{a,r} - d_{infectious} * IQ_{a,r}$$

$$IL'_{a,r} = l_{a,r} * d_{incubation} * IPS_{a,r} - d_{hosp} * IL_{a,r}$$

$$IH'_{a,r} = h_{a,r} * d_{incubation} * IPS_{a,r} - d_{hosp} * IH_{a,r}$$

$$R'_{a,r} = VE * d_{immune} * V_{a,r} + (1 - m1_r) * d_{infectious} * (IC_{a,r} + IQ_{a,r}) + (1 - m2_r) * d_{incubation} * IL_{a,r} + (1 - m3_r) * d_{hosp} * IH_{a,r}$$

$$D'_{a,r} = m1_r * d_{infectious} * (IC_{a,r} + IQ_{a,r}) + m2_r * d_{infectious} * IL_r + m3_r * d_{hosp} * IH_{a,r}$$

Some parameters are functions, as in the following.

$$hsp_a = Max[e^{\partial+\gamma(a)}, 0]$$

where $a$ is the age group and $\partial, \gamma$ are fitted parameters from an exponential regression.

$$h_{a,r} = e^{\log(\sigma)+\log\left(\frac{pr_{hosp}*hsp_a}{1-pr_{hosp}*hsp_a}\right)+\log\left(OR_{hosp}*(r-1)\right)}$$

where $r$ is the risk group index.

$$s_t = season * Cos\left[\left(t * \frac{2}{365} * \pi\right) + 0.1\right] + (1 - season)$$

Where $\pi$ is the numeric constant Pi, $t$ is the date, and $season$ controls the amplitude of the seasonal effect, larger numbers will increase the difference in transmission maximum and minimum values. The shift of 0.1 centres the minimum on the centre date between the hottest days of the year in Alberta's major cities, Edmonton and Calgary.

The contact matrix is decomposed into four locations: home, workplace, school, and other, where *other* comprises all remaining possible locations. Total contacts are given as the sum of contacts for each age to age ($_{a,k}$) entry in each of the matrices. Decomposing the total contact matrix by location allows for adjustments in contact rates at locations to align with policy interventions. For example, school closure is reflected by adjusting the school contact matrix ($school_{a,k,t}$) by a scalar value, for the duration of dates during which the school is closed:

$$school_{a,k,t} = iso_{school} * school_{a,k},$$

where $iso_{school} = 0$ while schools are closed, and $iso_{school} = 1$ otherwise (that is, $0 \leq iso_{school} \leq 1$).

Similarly, for interventions which would reduce contact rates in schools, $iso_{school,a}$ may be applied to specific age groups, adjusting the school contact matrix at various points in time according to age group.

Reductions $iso_{work}$, $iso_{home}$, and $iso_{other}$ are applied to the remaining contact matrices, which are then summed to generate the total matrix, $contact_{a,k}$, at each time step. Reduction in contacts for seniors age 75+ are applied directly to the total contact matrix, as a scalar multiplied through the rows and columns where *a* or *k* equals 16. Reduction in contacts for high-risk individuals are applied to the total contact matrix, only for those individuals whose *r* equals 2. For example:

$$\beta * \sum_{k=1}^{17} iso_{risk} * \frac{contact_{a,k}}{N_{k,\cdot}} [IPS_{k,\cdot} + IC_{k,\cdot} + IQH_{k,\cdot} + IL_{k,\cdot} + IH_{k,\cdot}] * S_{a,r}$$

where $iso_{risk} = 1$ for non-high-risk individuals.

#### 1.2 Assumptions

The CRAM makes the following assumptions:

- Only susceptible individuals who have not been exposed to COVID-19 are immunised. In practice, this may require serology testing prior to vaccine.
- The vaccine behaves as an all-or nothing vaccine, where the vaccine protects a proportion of those immunised corresponding to vaccine effectiveness.
- Daily contact rates are based on age group in CRAM. This limits the types of contacts represented in the model; for example, the model cannot represent cohorts of students (that is, students seeing the same limited group of students/staff each day).
- There are no additional school contacts between individuals older than 20 years in our model (for example, between school staff). Therefore, when contacts were increased with in-person schooling, increases in contact rates only applied to contacts between students and other students, and contacts between students and adults.
- All public health interventions were modelled by reducing the contacts of specific age or risk groups. This necessitates assumptions about contact reductions associated with each public health intervention (for example, how masks reduce contacts between school-aged children). Therefore, CRAM cannot test which public health interventions are most likely to reduce contacts; rather, it estimates the impact that reducing contacts through public health interventions may have on overall disease outcomes.
- Provincial level modelling is broadly informative at a local level. Where local case counts increase, this may indicate that additional actions need to be taken, including possible changes to schooling arrangements, but local factors relative to the provincial dynamics presented in these results should be taken into context.
- All strategies in place at the start of the period, other than vaccine and other clearly stated changes will remain in place for the duration of the model period. Therefore, the results reflect what might happen in a scenario where no mitigating actions are taken. Any interventions implemented at a local or regional level in response to an outbreak would result in a different number of expected infections and hospitalisations.

#### 1.3 Evaluating Minimum Iterations Required

Complex mathematical models such as the CRAM are computationally expensive and have long run times. To identify the sufficient number of simulations to achieve stable results, mean results for total number of cases, hospitalizations, total cost, and total utility loss were collected for simulation iterations from 10 to 800. The results from different simulation sizes were compared using Welches' t-test, which was selected since comparator samples can not be assumed to have equal variances due to inequal sample size.

The difference in means was compared for the four results (i.e., total number of cases, hospitalizations, total cost, and total utility loss) for simulations above a 'threshold' number of simulations. A rejection of the null hypothesis is interpreted as an indication that the number of simulations is insufficient. The number of iterations for which the null hypothesis is rejected above a given threshold is presented for different significance levels. Interpreting the 0.1 significance level as a low tolerance for differences in means, we found a minimum of 200 simulations is required to ensure stability in results. These values are presented in Table 1.

Table 1: Count of iterations by threshold for which difference in means statistically different than large-N simulation

| Simulation threshold | 0.01 significance | 0.05 significance | 0.1 significance |
|---|---|---|---|
| **50** | 0 | 36 | 61 |
| **75** | 0 | 22 | 36 |
| **100** | 0 | 4 | 11 |
| **150** | 0 | 0 | 0 |
| **200** | 0 | 0 | 0 |
| **250** | 0 | 0 | 0 |
| **300** | 0 | 0 | 0 |
| **350** | 0 | 0 | 0 |

### 2 Model Inputs

#### 2.1 Expert Elicitation

To generate estimates for parameters for which there was no information source, a rapid expert elicitation exercise was developed. The elicitation drew from the SHELF tool and adapted the questionnaire to be given through a web-based survey. Expert opinions were pooled to derive the median and 90% CI estimates for each value of interest.

An initial set of questions was disseminated to seven public health experts, identified in partnership with Alberta Health. Experts were provided with a short online training module, and responses were restricted to a 11-point Likert scale between 0 and 10. The median, and upper and lower tertiles were elicited, and beta distributions were fit to estimate response values. These values are presented in Table 2.

A second elicitation was disseminated to ten public health and education experts, using two return to school scenarios defined by the Alberta Government. Experts were provided with a webinar training, and then the questionnaire was disseminated online, with responses restricted to an 11-point Likert scale as before. The median, $5^{th}$ and $95^{th}$ quantiles were elicited, and beta distributions were fit to estimate response values. These values are presented in Table 3.

Note that following the expert elicitation, Alberta Health mandated mask use in schools. An additional reduction in contact rates using a uniform distribution ranging from 20-80% relative to the elicited values was applied to the school contact matrix. A uniform distribution was selected to account for the high degree of uncertainty in these estimates, meaning, for example, that 50% effectiveness is equally probably to 30% effectiveness. This value was based on a rapid review which found evidence of mask effectiveness at reducing spread, but uncertainty in the degree of effectiveness [1].

Table 2: Elicitation question and responses

| Quantity | Pooled response values, median [90% CI] | Elicited values and linear pool |
|---|---|---|
| Factor by which Albertans have reduced their contacts at home, given current public health orders and guidance | 0.586 [0.070, 0.995] | 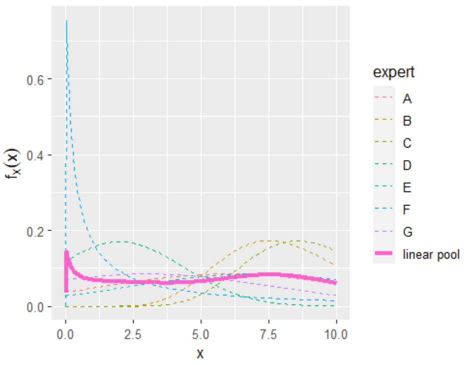 |
| Factor by which Albertans aged ≥75 years have reduced their contacts in all locations, given current public health orders and guidance | 0.387 [0.137, 0.945] | 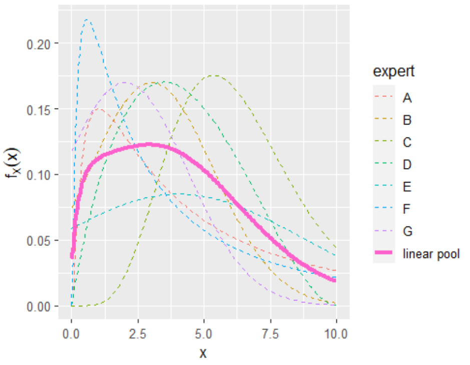 |
| Factor by which Albertans with a high-risk of severe outcomes have reduced their contacts in all locations, given current public health orders and guidance | 0.326 [0.065, 0.922] | 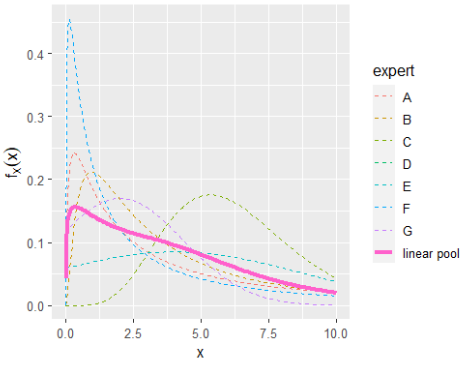 |

#### Table 3: School Elicitation question and responses

<table>
<tr><th>Quantity, Values</th><th>Age group: 5–9 years</th><th>Age group: 10–14 years</th><th>Age group: 15–19 years</th></tr>
<tr><td>
<table>
<tr><th>Age group</th><th>Contact reduction</th></tr>
<tr><td>5–9 years</td><td>0.692 [0.358, 0.869]</td></tr>
<tr><td>10–14 years</td><td>0.573 [0.314, 0.846]</td></tr>
<tr><td>15–19 years</td><td>0.522 [0.278, 0.861]</td></tr>
</table>
</td><td>
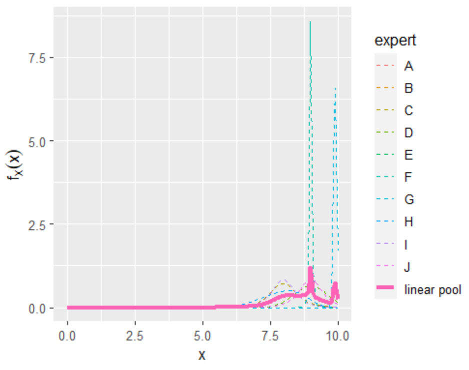

</td><td>
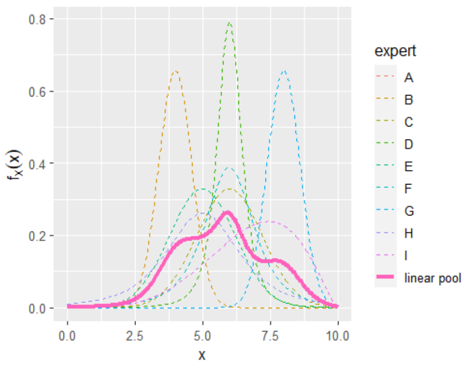

</td><td>
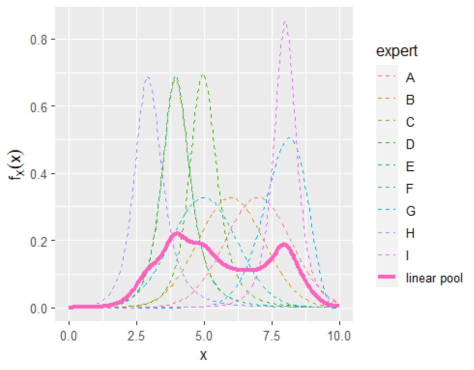

</td></tr>
<tr><td>
<table>
<tr><th>Age group</th><th>Proportion returning</th></tr>
<tr><td>5–9 years</td><td>0.760 [0.399, 0.933]</td></tr>
<tr><td>10–14 years</td><td>0.831 [0.399, 0.941]</td></tr>
<tr><td>15–19 years</td><td>0.860 [0.634, 0.994]</td></tr>
</table>
</td><td>
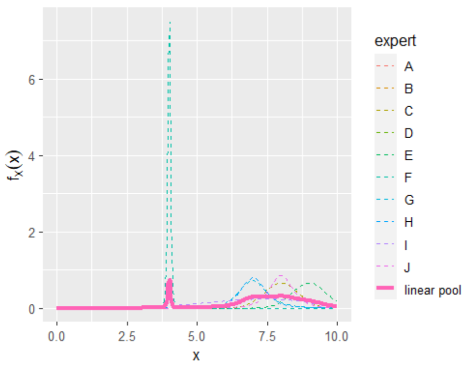

</td><td>
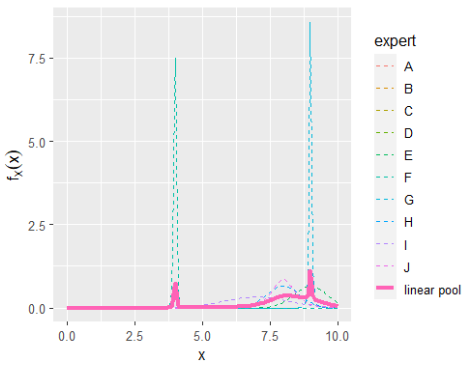

</td><td>
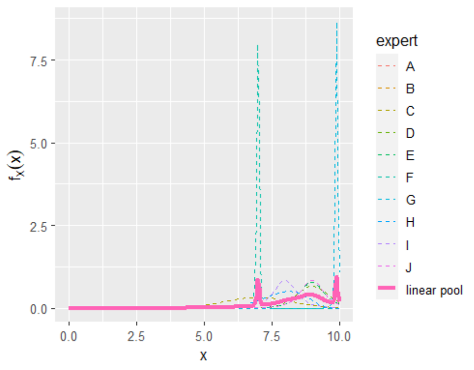

</td></tr>
</table>

CI: confidence interval

#### 2.2 Estimating Utility Decrement and Incremental Cost related to ‘Chronic COVID’

It is known that following SARS-Cov-19 infection, that some individuals experience long term health consequences, often referred to as ‘Chronic COVID’ or ‘Long COVID/Long-haulers’. To estimate the health utility decrement and incremental costs associated with Chronic COVID, we identified several chronic conditions which have emerged following SARS-Cov-19 infection: Chronic Fatigue, Diabetes, Chronic Kidney Disease, Chronic liver Disease, Adverse Cardiovascular events, and Psychiatric conditions. We converted incremental cost values identified in the literature to 2020 Canadian dollars following CADTH guidelines [2-10], and estimated utility decrements by identifying relevant general population comparison [11-17]. Estimates were combined by weighting relative to the prevalence of each condition in the pre-pandemic population [18-22]. Given that the duration of Chronic COVID is unknown, we conservatively include one year of utility decrement and incremental cost outcomes in our model inputs. Furthermore, we make the conservative assumption that Chronic COVID occurs only for hospitalized cases, and use the proportion of patients reporting ‘fatigue on discharge’ from [23], as chronic fatigue was a major contributor to utility decrements among the chronic conditions we identified. An excel workbook with full calculations, options to change the time period for discounting, and references is provided at https://github.com/eKirwin-IHE/chronicCOVID.
